# Liberté, Egalité, Fraternité… Contaminé? Estimating the impact of French municipal elections on COVID-19 spread in France

**DOI:** 10.1101/2020.06.24.20138990

**Authors:** Guilhem Cassan, Marc Sangnier

**Affiliations:** University of Namur, DEFIPP, CRED, CEPREMAP; University of Namur. Aix-Marseille University (Aix-Marseille School of Economics), CNRS, EHESS, Centrale Marseille

**Keywords:** COVID-19, Hospitalizations, Electoral turnout, Municipal elections, Prediction errors

## Abstract

On March 15, about 20, 000, 000 voters cast their vote for the first round of the 2020 French municipal elections. We investigate the extent to which this event contributed to the COVID-19 epidemics in France. To this end, we first predict each département’s own dynamics using information up to the election to calibrate a standard logistic model. We then take advantage of electoral turnout differences between départements to distinguish the impact of the election on prediction errors in hospitalizations from that of simultaneously implemented anti-contagion policies. We report a detrimental effect of the election in locations that were at relatively advanced stages of the epidemics by the time of the election. In contrast, we show that the election did not contribute to the epidemics in départements with lower infection levels by March 15. All in all, our estimates suggest that elections accounted for about 4, 000 excess hospitalizations by the end of March, which represents 15% of all hospitalizations by this time. They also suggest that holding elections in June may not be as detrimental.

## 1 Introduction

The two rounds of 2020 French municipal elections were planned to take place on March 15 and 22. By the beginning of March, the early spread of the COVID-19 epidemics led to a debate in the French society about whether the first round should actually be postponed. This option was finally rejected and Emmanuel Macron—the French President—announced on the evening of March 12 that the first round would take place as planned. This decision was accompanied by the announcement of the closing of all schools and universities by March 16 and was followed by an announcement by Edouard Philippe—the French Prime Minister—on March 14 about the closing of all non essential public spaces by the next day to prevent the spread of COVID 19. This marks the start of anti-contagion policies in France.

According to an Odoxa opinion poll published on March 12, 64% of French people approved the decision to maintain the election and 61% of voters reported that the epidemics won’t change their decision to vote. On March 15, 19, 863, 660 out of 44, 650, 472 voters in metropolitan France cast their vote, with no alternative but to go to the voting booth in order to do so. On March 16, Emmanuel Macron announced that strict lockdown measures would be put in place from March 17 onwards and that the second round of the municipal elections was postponed *sine die*.

In this paper, we show that the first round of 2020 municipal elections caused an acceleration of the COVID-19 epidemics in metropolitan France. Our estimates suggest that elections accounted for about 4, 000 excess hospitalizations by the end of March, which represents 15% of all hospitalizations by this time.

Our methodology takes advantage of electoral turnout differences between *départements*— the third highest administrative level—to distinguish the impact of the election on hospitalizations from that of simultaneously implemented anti-contagion policies. Our approach builds on methods from the abnormal financial returns and public policies evaluation literatures (see MacKinlay 1997, Duflo 2001, Fisman 2001, Guidolin and La Ferrara 2007, DellaVigna and Ferrara 2010, Coulomb and Sangnier 2014 and Cassan 2019 among others). We proceed in two steps. First, we fit for each département a simple epidemic model of hospitalizations for COVID-19 suspicion on the period that excludes hospitalizations that might relate to events that took place by March 15 or in the following days. We then use these models to predict the evolution of the epidemics in each département as if propagation conditions were held constant and compute daily predictions errors as the difference between the realized and predicted cumulated number of hospitalizations in each département.

Second, we relate prediction errors to March 15 turnout and to differences in the epidemics stage across départements by that date. This approach allows us to asses the causal effect of elections on hospitalizations while accounting for other contemporaneous events such as anti-contagion policies which were *a priori* uniform throughout the country. It explicitly accounts for different dynamics at the local level and builds on the assumption that prediction errors should not be related to turnout and March 15 epidemics stage in the absence of an effect of the election on hospitalizations. We show that post-calibration errors are increasing with turnout in départements where the COVID-19 epidemics was active by the day of the election. In contrast, turnout is not related to post-calibration errors in locations with low COVID-19 activity by March 15.

Our identification strategy is akin to a quadruple-differences method, effectively taking advantage of the following differences: (i) the within-département difference between realized and predicted hospitalizations; (ii) the within-département difference between periods before and after the election; (iii) the between-départements difference in electoral turnout; and (iv) the between-départements difference in epidemics intensity on the election day. This combinations of differences allows us to asses the causal impact of the elections on hospitalizations for COVID-19 suspicion. Importantly, our estimation strategy also allows us to explicitly account for other factors that might explain differences in the dynamics of the epidemics, such as population density or the share of elderly population. This approach allows us to quantify the causal impact of the elections on COVID-19 related hospitalization for metropolitan France. Moreover it also allows us to discuss the likely impact of the second round of the elections on the epidemic, which we (tentatively) anticipate to be non statistically detectable.

As highlighted by Hsiang et al. (2020), most studies that analyze the impact of policies on COVID-19 rely on complex epidemiological models which require a detailed knowledge of the fundamental epidemiological parameters of COVID-19. Our approach, taken from the standard methods of reduced form econometrics commonly used to assess the impact of public polices (Angrist and Pischke 2009), does not require such detailed information. Our approach allows us to disentangle the impact of the election from others confounding shocks that may have hidden it without requiring much information about the specific mechanism of the disease itself.

To the best of our knowledge, only two other papers attempt to evaluate the impact of 2020 municipal elections on the COVID-19 epidemics in France. Zeitoun et al. (2020) compare the post-election epidemic trajectories of départements with high and low turnout as well as high and low intensity of the epidemics. They report no impact of the elections on the spread of the epidemic in France. We depart from this study by explicitly allowing each département to follow a specific epidemic dynamics. Bertoli et al. (2020) study the effect of the municipal elections on excess mortality at home in the subset of French municipalities that have no hospital. Using an instrumental variable approach to predict turnout at the very local level, they report a qualitatively strong impact of the election on excess mortality. We depart from this study by focusing on départements—an observation unit that is *a priori* relevant to study the epidemics’ dynamics—rather than municipalities, by using data that cover the whole metropolitan France, and by implementing a methodology that allow us to provide a quantitative estimate of the impact of the elections on the epidemics.

## 2 Data and methodology

This section presents the data used in this paper as well as the methodology we rely on to assess the impact of the March 15 elections on the spread of the COVID-19 epidemics.

### 2.1 Data

Our analysis relies on two main datasets: hospitalizations for COVID-19 suspicion and electoral turnout at the 2020 French municipal elections. We also make use of demographic data at the département level.

Hospitalization data are open access data published by the French government. Data are based on hospitals’ reports and present the daily counts of hospitalization decisions for COVID-19 suspicion at département level from February 24 onwards. 2020 electoral turnout data for the first round of municipal elections are official electoral records available at the city-level. We aggregated these data at the département level. We proceed identically with 2014 electoral turnout data. Finally, we collect official total population and population aged above 60 on January 1, 2020 in each département from official records and construct population density at the département level using départements area information.

### 2.2 Methodology

We use the daily cumulated number of hospitalizations for COVID-19 suspicion to fit a series of département-level epidemics trajectories up to the date at which individuals contaminated on March 15 start being hospitalized. We separately estimate the following standard logistic model of epidemiological trajectory for each département *d*:

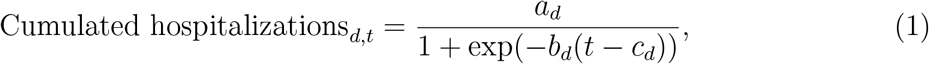

where *a*_*d*_, *b*_*d*_ and *c*_*d*_ capture the asymptotic level, the inflection date and the scale of the epidemics trajectory in département *d*, respectively. We estimate equation (1) using all dates *t* until March 26, i.e. 11 days after the elections took place. This 11-day lag is one day shorter than the median estimate of the number of days from infection to hospitalization suggested by the clinical studies literature.^1^ As a result, the model’s forecasts can be interpreted as départements trajectories in the absence of any event that took place since March 15.

We estimated model (1) for each of the 96 départements of metropolitan France. The model was successfully estimated for 91 départements. The 5 départements for which we are not able to calibrate the model are départements that do not exhibit sufficient variation in hospitalizations until March 26 to allow for parameters’ estimation. These départements account for 1.6% of the total French population.

Following insights from the literature on short term epidemiological forecast (Chowell et al., 2019; Roosa et al., 2020a,b), we use the series of estimated parameters *â*_*d*_,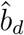 and *ĉ*_*d*_ to predict for each département the daily cumulated number of hospitalizations up to 7 days after the end of the calibration period, i.e. up to April 2. Predicted trajectories proxy the evolution of the epidemics in each département in the absence of the election and of any other shock contemporary or posterior to the election, such as lockdown policies. We use the actual number of hospitalizations for COVID-19 suspicion in each département to construct prediction errors in hospitalizations per 100, 000 inhabitant. As shown by Figures S1(a) and (b) available from Supplementary material A, predictions errors are generally positive over the post-calibration period, which suggest that most départements surpass their predicted epidemics trajectories after March 15. Our interest is however not to assess whether it is possible to correctly predict the evolution of the epidemics, nor to estimate whether policies implemented after this date were able to twist trajectories.

In contrast, our interest lies in whether deviations in epidemics trajectories depend on the March 15 elections. To this end, we take advantage of two sources of variations to assess whether this event impacted the spread of the COVID-19 epidemics. First, we distinguish between départements depending on the local stage of the epidemics by the time of the election. Second, we use differences in electoral turnout to proxy for difference in exposure across départements at comparable stages of the epidemics. We then estimate the following expression:

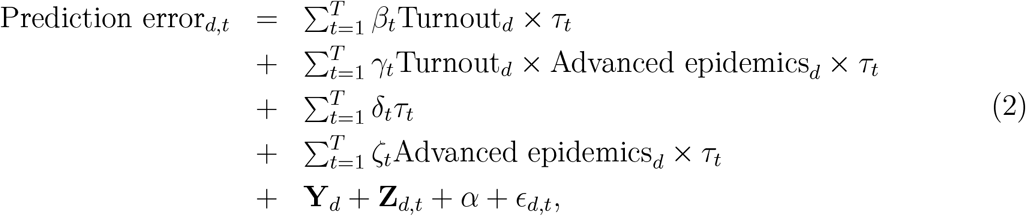

where Prediction error_*d,t*_ is the difference between actual and predicted cumulated hospitalizations per 100, 000 inhabitants in département *d* on day *t*, Turnout_*d*_ is electoral turnout on March 15 in département *d, τ*_*t*_ is a variable equal to 1 on day *t*, Advanced epidemics_*d*_ is a variable equal to 1 for départements at advanced stages of the epidemics on March 15, series of *δ* and *ζ* coefficients account for daily patterns in prediction errors across départements in both groups, **Y**_*d*_ is a vector of département fixed effects which account for département-specific patterns, **Z**_*dt*_ is a vector of interactions between day fixed effects and départements population density and share of population aged above 60, *α* is a constant term, and *E*_*dt*_ is the error term. We estimate expression (2) using ordinary least squares and cluster standard errors at the départment and day levels. The sample is made of all days from March 1 to April 2, 2020.

Consistent with the aforementioned 11-day lag between infection and hospitalization, we use cumulated hospitalizations per 100,000 inhabitants on March 26 to construct the Advanced epidemics_*d*_ variable that distinguishes between départements depending on the stage of the epidemics by the day of the election. We arbitrarily distinguish between départments in the bottom third of the COVID-19 epidemics according to this measure and others.^2^ The latter are considered as locations at relatively more advanced stages of the epidemics.^3^

In expression (2), the main parameters of interest are the estimated series of *β*_*t*_ and *γ*_*t*_. These coefficients indicate the impact of electoral turnout on hospitalizations for départements with low infection and its additional effect for départements with high infection, respectively. Under the assumption that the March 15 elections impacted epidemics trajectories only in locations that were at advanced stages of the epidemics by that day, we expect *β*s to be close to zero and *γ*s to be positive in the post-calibration period.

Supplementary material E displays results obtained using alternative definitions of the group of départements considered as at advanced epidemic stage by the day of the election. Supplementary material F presents point estimates obtained when removing each département one-by-one.

### 2.3 Threat to identification

A key assumption for the above presented approach to allow us to safely assess the impact of municipal elections on the dynamics of the COVID-19 epidemics is that electoral turnout on March 15 is unrelated to the stage of the epidemics by that date. Namely, turnout was low as only 45% of voters cast their vote, compared to 64% at the 2014 municipal elections. There is a wide consensus in the French society that this low turnout was mainly caused by the fear of contagion. This might actually be the case but would be a threat to identification only if differences in turnout across départements ended up being related to differences in the epidemics across départements. We find no evidence of such a correlation between the level of turnout in a départment and the information on the spread of the epidemic in that département on the day of the election. This is best illustrated by Figure 1(a) which plots turnout against publicly known cumulated hospitalizations on March 15. Turnout appears evenly distributed at each stage of the epidemics.

**Figure 1:**
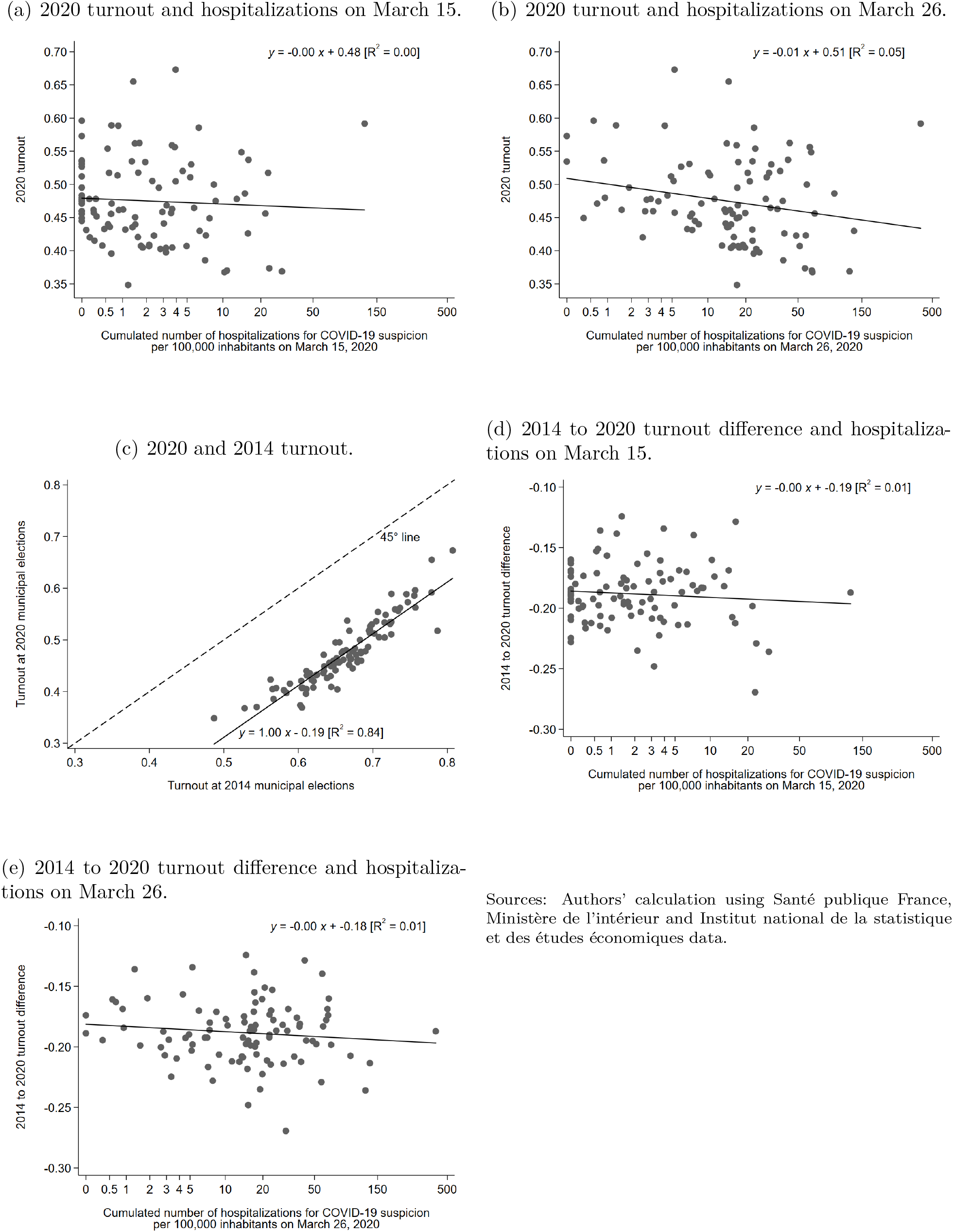
Electoral turnout and the COVID-19 epidemics.

Figure 1(b) further accounts for the 11-day lag from infection to hospitalization to better capture the underlying stage of the epidemics in each département and only reveals a weakly decreasing link between turnout and hospitalizations. In contrast, turnout at the 2020 municipal elections is strongly correlated with turnout at the preceding municipal elections that took place in 2014 as shown by Figure 1(c). This Figure shows that the shift in turnout was uniform across départements. Figure 1(d) and (e) further illustrate this claim by plotting the 2014 to 2020 turnout difference against cumulated hospitalizations on March 15 and 26, respectively.

All in all, while the COVID-19 epidemics might have impacted turnout at the 2020 municipal elections—a question that is beyond the scope of this paper—differences in the spread of the epidemics by March 15 did not translate into differences in turnout across départements, thereby allowing us to confidently interpret estimates that will be delivered by our identification strategy.

### 2.4 Discussion of the methodology

The outcome of interest of our approach is the extent to which the first-step predictive model fails to predict the evolution of hospitalizations. In the absence of an effect of the election on hospitalization, our model should make similar errors of predictions across départements, no matter their turnout.

However, if the election indeed had an effect on the epidemic, the prediction errors should be relatively larger in départements with relatively higher turnout. Indeed, if elections did contribute to spread the epidemics, the predictive model should underestimate by a larger amount the number of cases in départements with high turnout compared to départements with low electoral turnout. And this stronger underestimation should start only when individuals infected on the election day are hospitalized, not before. Similarly, the effect of turnout on the epidemics’ spread should only exist in départements in which the contagious individuals are indeed present: a high turnout in a département with no or few contagious individuals should result in 0 additional contagions.

We therefore analyse prediction errors via a triple-difference approach: not only do we compare départements with high and low turnout before and after the elections, but we study how this double-difference varies between départements with very low infection rates around the election date and other départements. We would expect turnout to only have an effect on the epidemics in départements already affected by the epidemics at the time of the election.

This approach has several advantages. First, it does not requires blind faith in the ability of the predictive model to deliver accurate predictions. In fact, it does rely on the model’s predictions being wrong while *a priori* uncorrelated with turnout under the null assumption that elections had no impact on the spread of the COVID-19 epidemics. Second, the event study aspect of the approach allows us to exactly observe when the prediction errors become correlated with turnout: predictions error should start being correlated with turnout *only* when people infected on the election day start showing up at hospital, that is, only when enough (but not too much) time has passed since the election for the symptoms to be severe enough to lead to hospitalization. This approach therefore automatically implements a sanity check as the correlation between the model’s prediction errors and turnout should emerge with a lag compared to the election date, but not too long a lag.

A drawback of our approach is however that these type of simple predictive models are typically precise in the short run only, so that predictions are likely to become more and more noisy the further away we move from the end of the calibration period, which should result in imprecise estimates. This is the reason why we stop the analysis 7 days after the end of the model’s fit. This time span is however likely to cover most of the additional hospitalizations that could be related to the March 15 elections as severe lockdown policies were implemented in the days that immediately follow, thereby limiting further transmission by people who would have been contaminated on that day.

## 3 Results

This section presents and interprets the results of the study.

### 3.1 Relationship between electoral turnout and hospitalizations

Figure 2 presents the series of *β*_*t*_ and *γ*_*t*_ coefficients estimated from equation 2. The series of *β*_*t*_ coefficients stays small and insignificant over both the calibration and prediction periods. This shows that turnout did not have any impact on hospitalizations in départements with very low infection rates on the day of the election. Similarly, the series of *γ*_*t*_ coefficients is close to zero and statistically insignificant over the calibration period. In contrast, the series starts increasing by March 27. This suggests that turnout is positively associated with hospitalizations in départements in which there were a relatively high number of contagious individuals by the election day exactly 12 days after the day of the election, in line with the 12-day lag between infection and hospitalization estimated by the literature.

**Figure 2:**
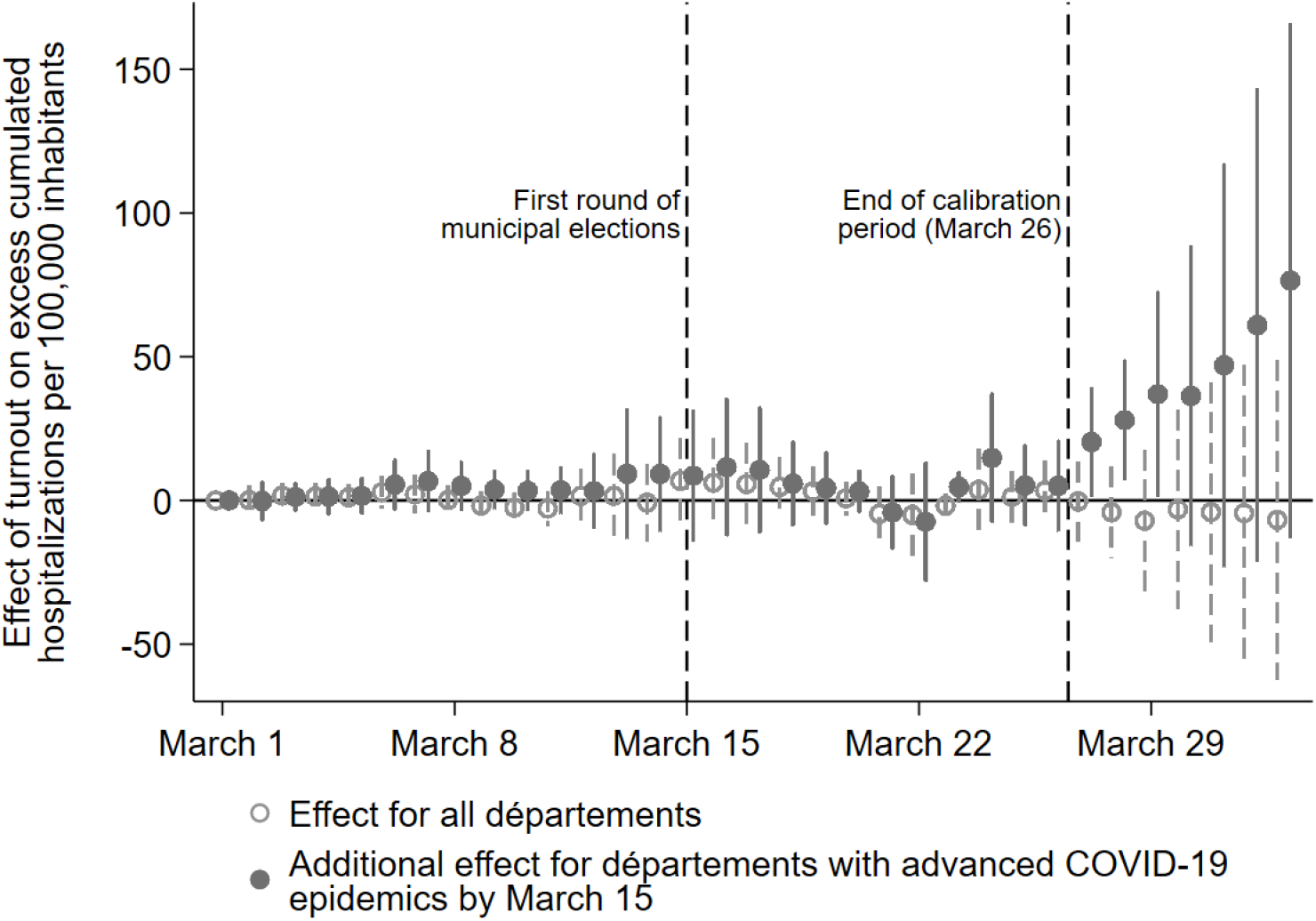
Relationship between electoral turnout and excess hospitalizations. Estimates of *β_t_* and *γ_t_* from equation (2) (see section 2). Vertical lines are 95% confidence intervals. *Département with advanced COVID-19 epidemics by March 15* are départements in the top two thirds of the distribution of cumulated hospitalizations for COVID-19 suspicion by March 26.

As discussed in section 2, the uncovered positive relationship can be interpreted as evidence of a causal relationship from the election to hospitalizations. However, beyond the increasing pattern of the series of *γ*_*t*_ coefficients after March 27, Figure 2 also displays increasing standard errors of the estimates as close as 3 days from the end of the calibration period. This feature calls for caution in the interpretation of the point estimates.

### 3.2 Quantification of the total effect

As shown by Table S1 available from Supplementary material C, *γ* coefficients estimated for March 27, 28 and 29 correspond to 20.3 (p-value = 0.032), 27.9 (0.009) and 37.0 (0.041) excess cumulated hospitalizations, respectively, for an hypothetical change in turnout from 0% to 100% in départements at relatively advanced stages of the epidemics by the election day. This contrasts with the coefficient estimated on the following days that are much larger and less precisely estimated. For instance, coefficients estimated for April 1 and 2 correspond to 61.0 (p-value = 0.139) and 76.5 (0.090) excess cumulated hospitalizations, respectively, for the same hypothetical change.

Actual electoral turnout data can help us to quantify the contribution of the March 15 elections to the COVID-19 epidemics. To this end, we use estimated coefficients of equation (2) and compute turnout-related excess cumulated hospitalizations per 100,000 inhabitants on each day from the end of the calibration period to April 2 in départements that were at advanced stages of the epidemics on March 15 as:

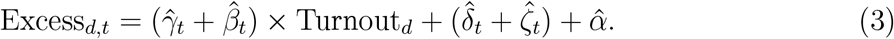

We then multiply these figures by each département population to obtain absolute figures and set excess hospitalizations to zero in départements with low COVID-19 activity on the election day. Figure 3 plots elections-related excess and actual cumulated hospitalizations at the national level. Our estimates suggest that the March 15 municipal elections accounted for 1, 811, 1, 591 and 2, 860 hospitalizations on March 27, 28 and 29, respectively. These figures represent 9.7%, 7.7% and 12.8% of cumulated hospitalizations by these days, respectively. Less reliable estimates available for April 1 and 2 suggest that elections accounted for 7, 884 and 9, 496 cumulated hospitalizations by these days, representing 27.9% and 31.7% of actual hospitalizations at these dates. These latter estimates are presumably less solid because of larger prediction and estimation errors as we move away from the end if the calibration period. As such, they must be considered as upper bounds. In contrast, March 27–31 estimates suggests that elections account for about 4, 000 hospitalizations, which represents 15% of all cumulated hospitalizations for COVID-19 suspicion in metropolitan France by March 31.

**Figure 3:**
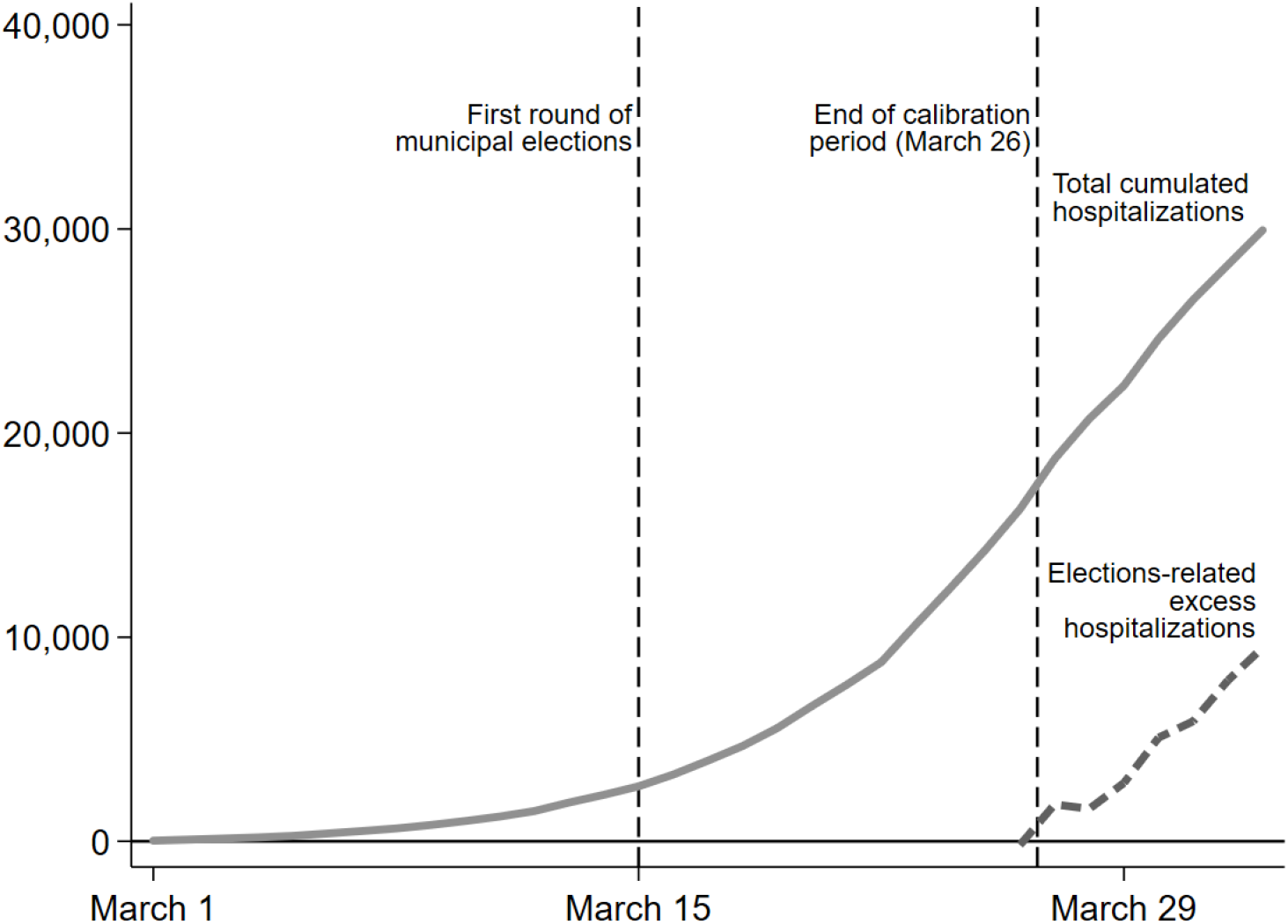
Election-related excess hospitalizations. Estimated election-related excess hospitalizations are computed using equation (3). See the text for more details.

## 4 Conclusion

According to our analysis, measures implemented on March 15 to prevent contamination in voting stations by the first round of the 2020 French municipal elections were not fully effective and resulted in about 4, 000 additional hospitalizations by the end of March. That is, 15% of hospitalizations accumulated by that time can be considered as direct consequences of the election.

On May 22, the French government announced that the second round of the municipal elections will take place on June 28 in municipalities in which no list gained majority in the first round. About 16, 000, 000 voters—mostly in the largest municipalities—are called to vote again. In the more distant future, départemental and regional elections are scheduled for 2021. For sure, anti-contagion policies implemented on March 15 need to be reviewed and improved to prevent these (and future) elections from spreading the current or future epidemics. Still, this study’s results allow us to qualitatively evaluate the likelihood that the 28 June second round will trigger a new COVID-19 wave.

After more than two months of lockdown and severe anti-contagion policies, the epidemic situation in June is not comparable to that in March. While the first round took place at the beginning of the exponential part of epidemics curve, the lockdown essentially amounted to a reset of infections. Infection levels are thus likely to be much lower in June than they were on March 15.

According to our estimates, départements in the bottom third of the distribution of cumulated hospitalizations per 100, 000 inhabitants by March 26 did not experience any worsening of their epidemic trajectory because of the March 15 election. This arbitrary threshold corresponds to 14 cumulated hospitalizations per 100, 000 inhabitants in the distribution constructed using hospitalization records since February 24, i.e, 32 days to March 26.

To asses the situation of French départements *vis-à-vis* an election to be organised in June, we use the latest hospitalizations record available by June 19, 2020 and reconstruct the 32-day cumulated number of hospitalizations per 100,000 inhabitants by that date in each département. As shown by Figure 4, 81 out of 96 départements experience lower than 14 reconstructed cumulated hospitalizations by June 19. This suggests that these départements would have been locations where an election won’t have accelerated the situation if organized 11 days before, i.e, by June 8.

**Figure 4:**
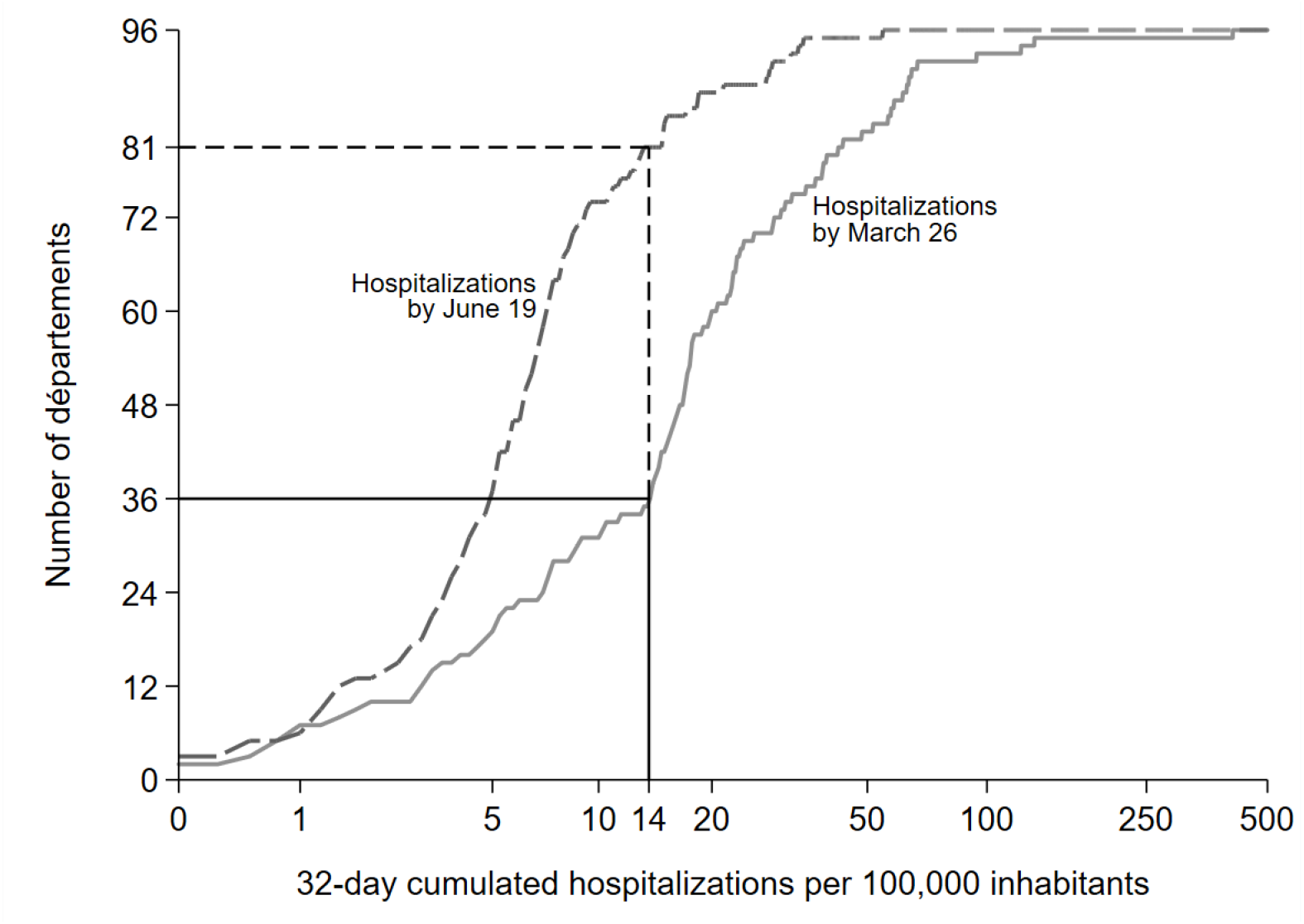
32-day cumulated hospitalizations per 100, 000 inhabitants on March 26 and June 19. 96 départements of metropolitan France. Distributions of hospitalizations per 100, 000 inhabitants cumulated over 32 days until March 26 and June 19, 2020. The vertical line at 14 hospitalizations per 100, 000 inhabitants correspond to the bottom third of the distribution for March 26 when excluding the 5 départements for which model (1) cannot be calibrated because of insufficient variation in hospitalizations until March 26.

It is important to keep in mind that (i) the threshold we use has no medical content and should only be understood as a way to compare two situations, and that (ii) our results, and thus our forecasts about end of June 2020, rely on estimates that are average effects by construction and might thus encompass heterogeneous situations. These limits in mind, it is likely that additional départements will cross the above mentioned threshold from here to June 28 if the epidemiological situation continues to improve. As a consequence, it is likely that holding elections on June 28 will not cause a statistically detectable number of new contaminations.

## Data Availability

All data used in this study are publicly available. Complete replication files are available from the authors' webpages.

http://marcsangnier.free.fr/divers/Liberte_Egalite_Fraternite_Contamine_2020_06_22_replication.7z

https://perso.unamur.be/~gcassan/stuff/Liberte_Egalite_Fraternite_Contamine_2020_06_22_replication.7z

## Supplementary material

### A Prediction Errors

**Figure S1:**
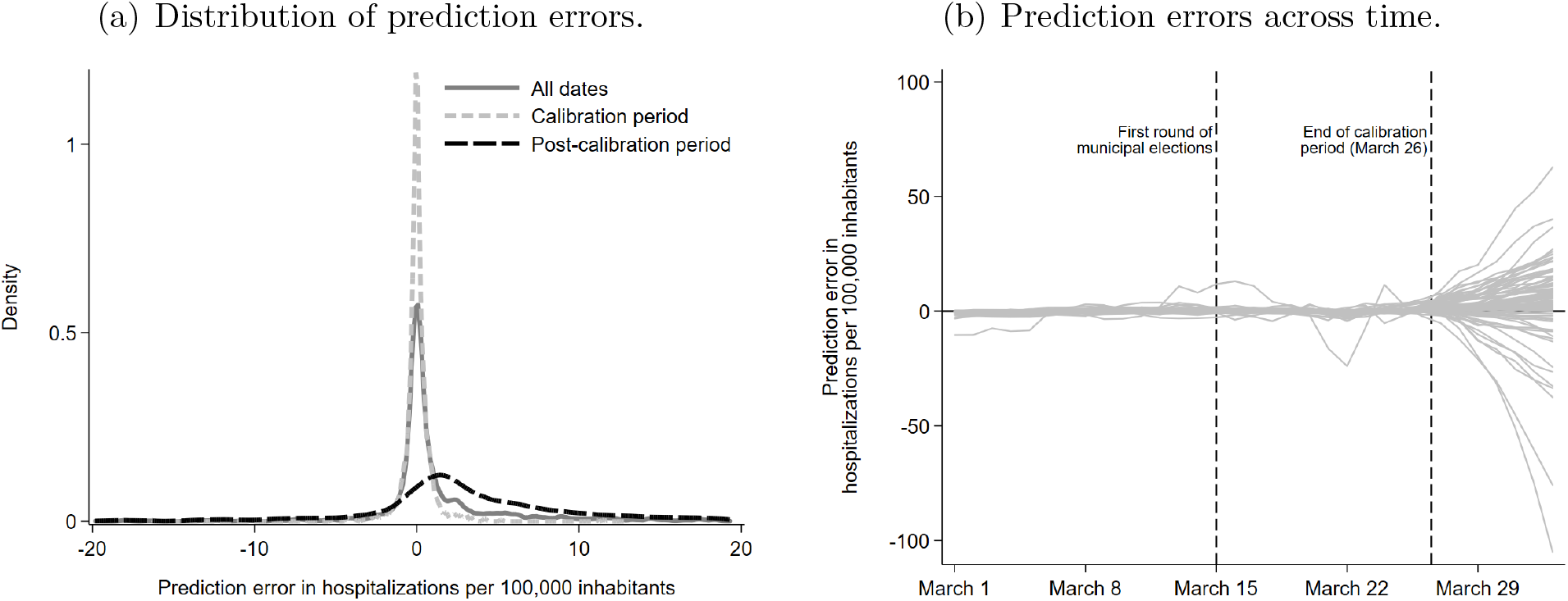
Prediction errors. Figures (a) and (b) plot the prediction errors of model (1) calibrated until March 26. Predictions are computed up to 7 after the end of the calibration period. See section 2 for more details. Figure (a) excludes prediction errors out of the [*−*20, 20] range.

### B Cumulated number of hospitalizations per 100,000 inhabitants on March 26

**Figure S2:**
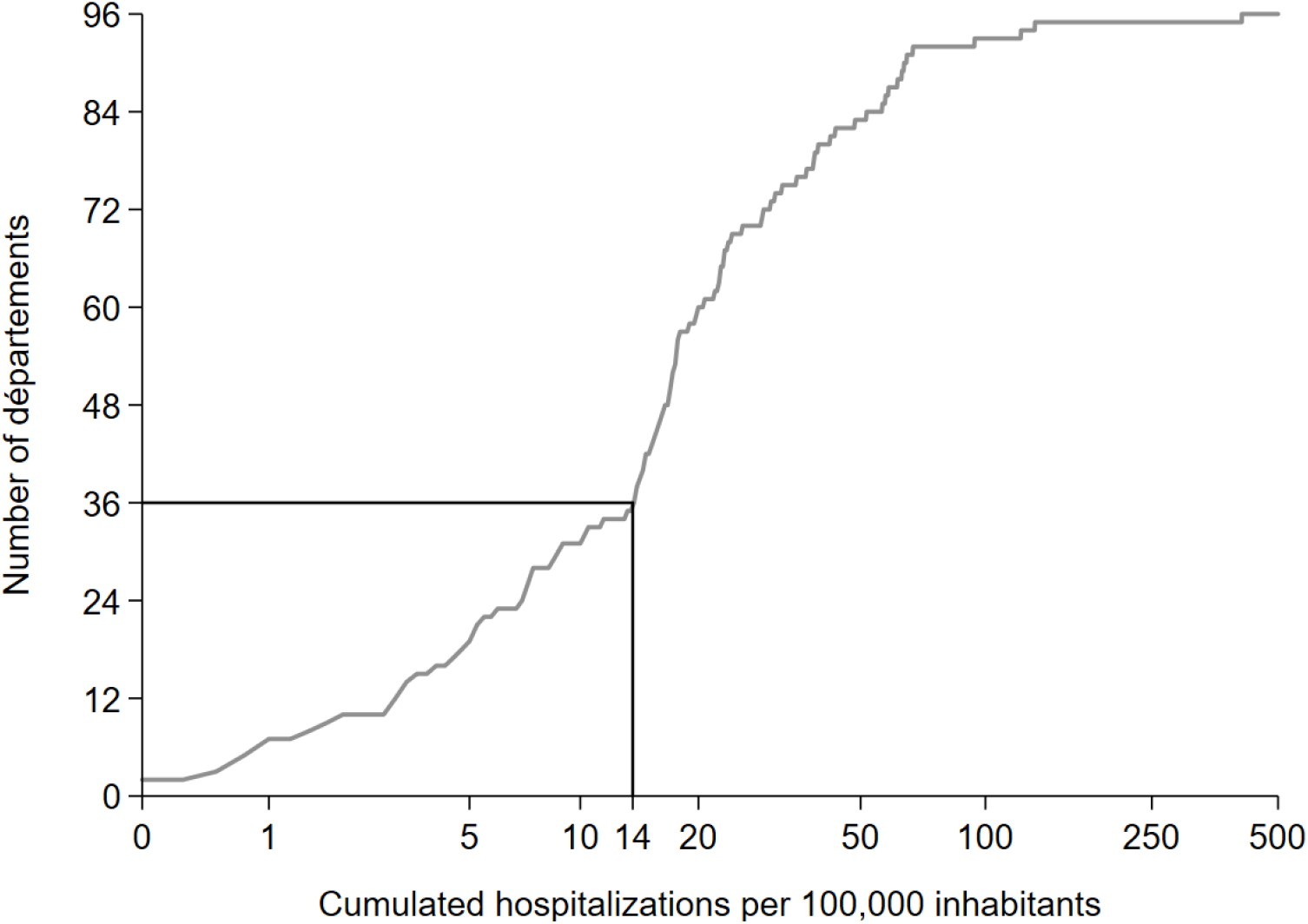
Distribution of cumulated hospitalizations per 100, 000 inhabitants on March 26. 96 départements of metropolitan France. The vertical line at 14 hospitalizations per 100, 000 inhabitants correspond to the bottom third of the distribution when excluding the 5 départements for which model (1) cannot be calibrated because of insufficient variation in hospitalizations until March 26.

### C Detailed regression results

**Table S1:** Estimates of the effect of turnout on excess hospitalizations.

|  | Coefficient (standard error) [p-value] |  |
| --- | --- | --- |
|  | Effect of turnout | Additional effect of turnout for départements with advanced epidemics |
| March 25 | 1.228<br>(4.360)<br>[0.780] | 5.209<br>(6.730)<br>[0.445] |
| March 26 | 3.376<br>(5.030)<br>[0.507] | 5.077<br>(7.566)<br>[0.507] |
| March 27 | -0.316<br>(6.809)<br>[0.963] | 20.333<br>(9.083)<br>[0.032] |
| March 28 | -4.135<br>(7.796)<br>[0.599] | 27.940<br>(10.080)<br>[0.009] |
| March 29 | -7.078<br>(12.094)<br>[0.562] | 37.020<br>(17.341)<br>[0.041] |
| March 30 | -3.117<br>(17.049)<br>[0.856] | 36.360<br>(25.434)<br>[0.163] |
| March 31 | -4.044<br>(22.145)<br>[0.856] | 47.031<br>(34.232)<br>[0.179] |
| April 1 | -4.416<br>(25.192)<br>[0.862] | 60.980<br>(40.144)<br>[0.139] |
| April 2 | -6.745<br>(27.230)<br>[0.806] | 76.521<br>(43.732)<br>[0.090] |

### D Results using 10 days as time from infection to hospitalization

Figure S3 displays results obtained using 10 days, rather than 11 days, as lag from infection to hospitalization. March 25 is thus used in lieu of March 26 as the date at which the calibration period ends and as the day at which we distinguish between départements with low or high epidemics by the time of the municipal elections. As the prediction model is calibrated on a shorter period, model (1) is successfully estimated for only 88 out of the 96 départements. The 8 left-aside départements account for 4.0% of the French population.

Figure S3(a) presents coefficients of interest when estimating equation (2) using March 25 both as the end of the calibration period for model (1) and as the date at which the categorization between high and low infection départements is done using the bottom third of the distribution of hospitalizations per 100, 000 inhabitants. Although less precisely estimated, the patterns of coefficient over the days after the end of the calibration period is similar to that found using March 16. Figure S3(b) displays the corresponding total excess hospitalizations associated with the elections.

**Figure S3:**
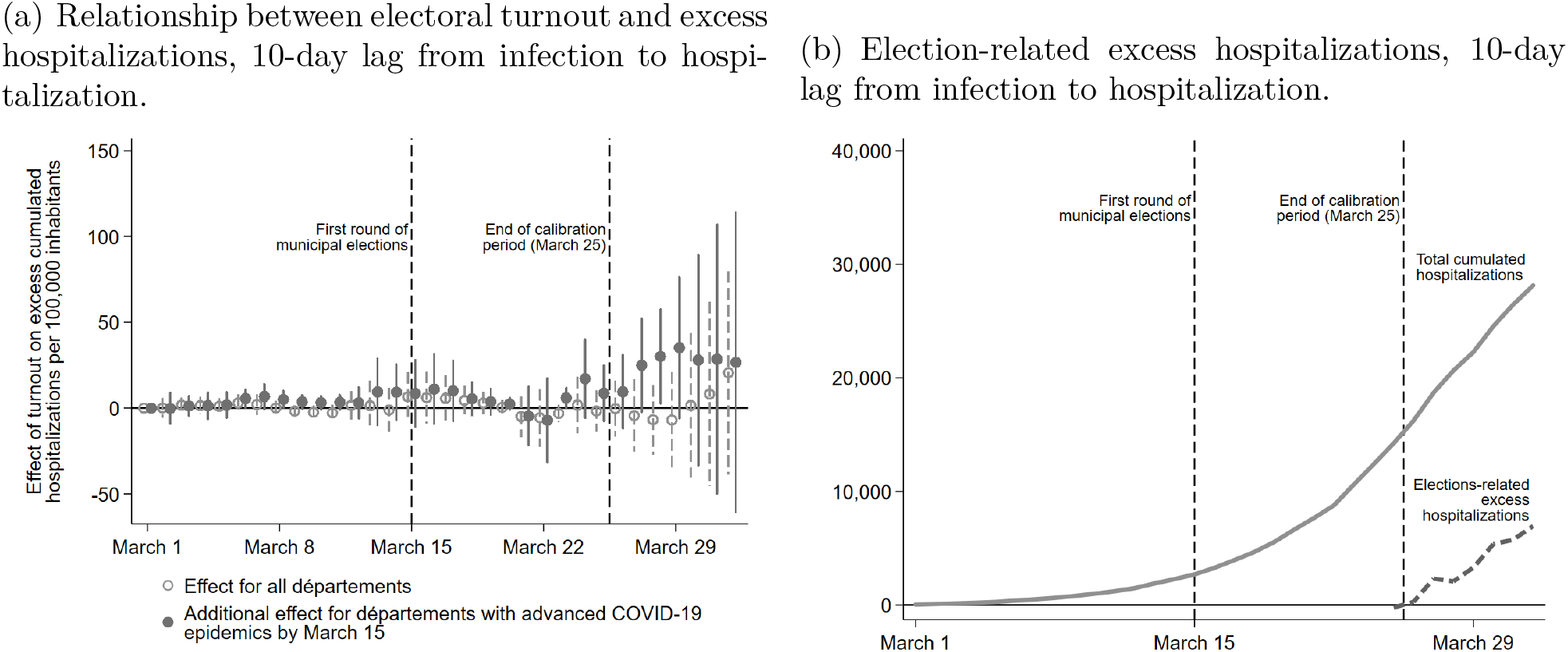
Estimates using 10 days as time from infection to hospitalization. Figures (a) and (b) mimic Figures 2 and 3 from the main text but use March 25 in lieu of March 26. Vertical lines are 95% confidence intervals.

### E Alternative definitions of advanced epidemic stage

Figure S4(a) presents the estimated coefficients of equation (2), using the 25th percentile of the distribution of cumulated hospitalizations per 100,000 inhabitants across départements on March 26 to construct the group of départements considered as at advanced stage of the epidemics by March 15. Figure S4(b) plots the corresponding total excess hospitalizations associated with the elections. Figure S4(c) and (d) further display results obtained when identifying départements with advanced epidemics activity as départements which that experienced more than 7 days of increase in hospitalization until March 26.

**Figure S4:**
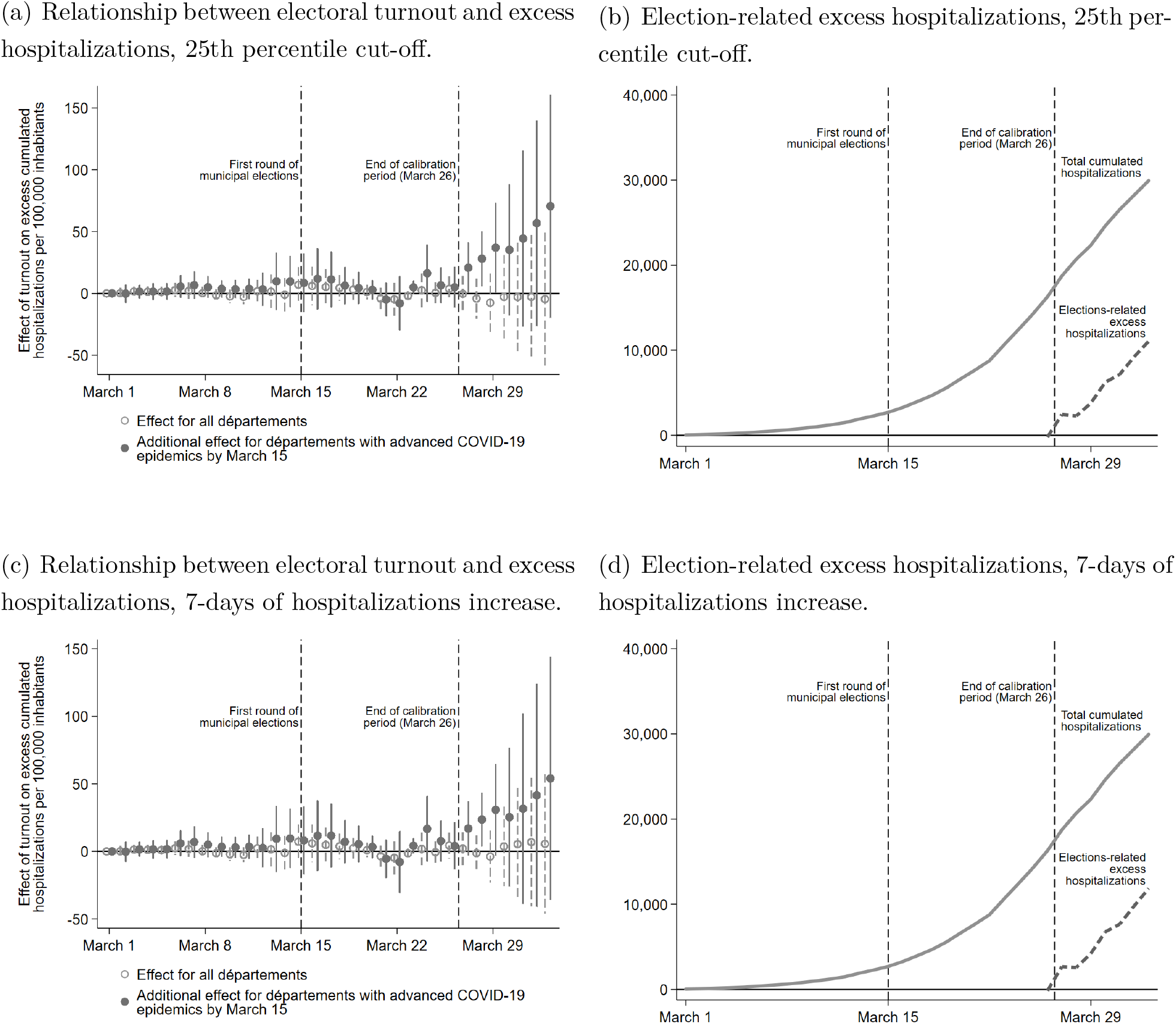
Estimates using alternative definitions of advanced epidemic stage by March 15. Figures (a) and (c) mimic Figure 2 from the main text. Vertical lines are 95% confidence intervals. Figures (b) and (d) mimic Figure 3 from the main text.

### F Results removing départements one-by-one

Figure S5 reproduces the results of Regression 2, but omitting each département one by one, to check if an outlier département is not driving all results. It can be seen that this is not the case.

**Figure S5:**
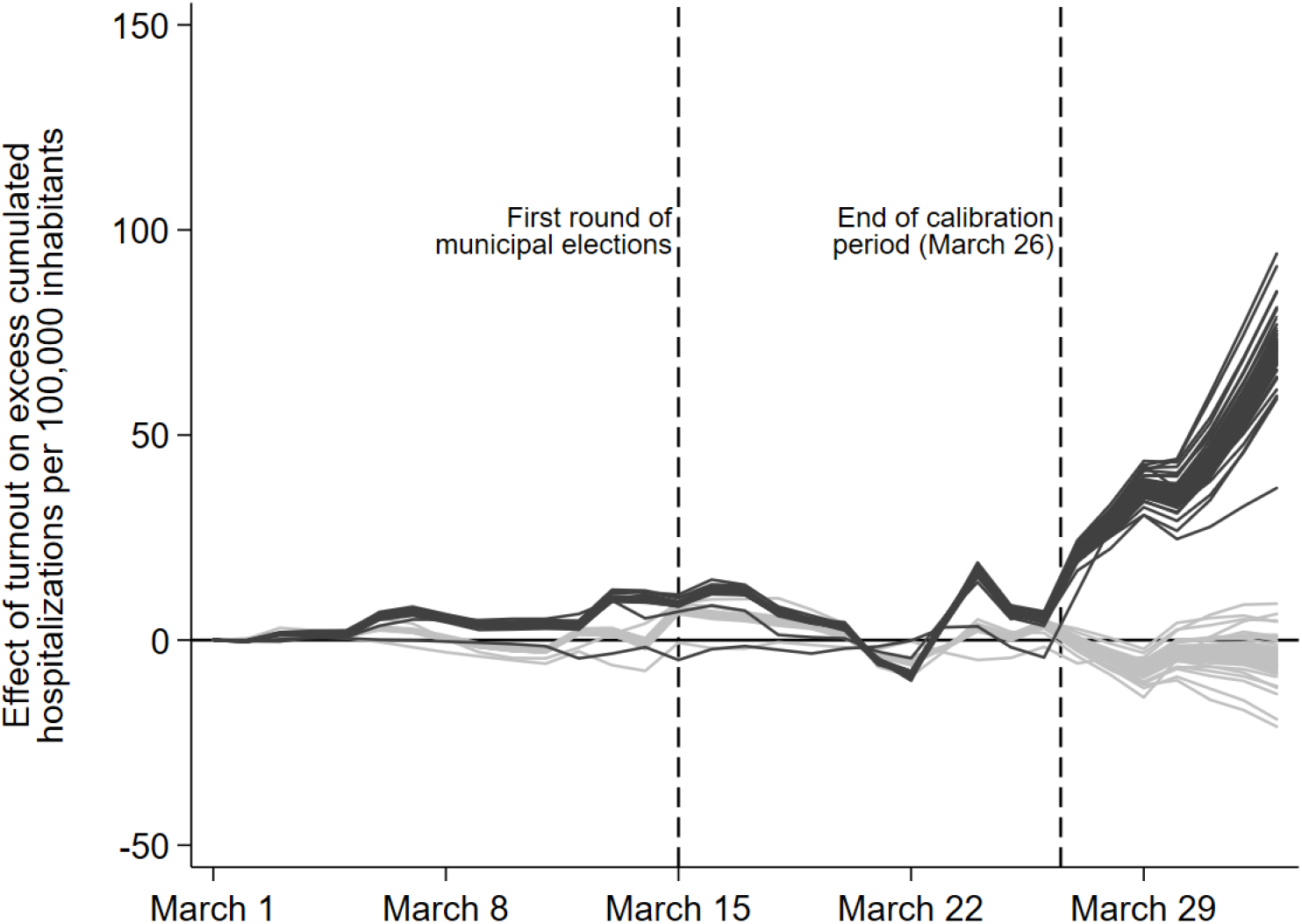
Relationship between electoral turnout and excess hospitalizations. Series of estimates of *β_t_* and *γ_t_* from equation (2) (see section 2). Each line corresponds to a series of coefficients obtained when excluding a given departement from the sample. *Département with advanced COVID-19 epidemics by March 15* are départements in the top two thirds of the distribution of cumulated hospitalizations for COVID-19 suspicion by March 26.

## Footnotes

1 Using Chinese data, Li et al. (2020); Chan et al. (2020); Guan et al. (2020) estimate that the time from infection to onset of symptoms is between 4 and 5 days, while Li et al. (2020), Huang et al. (2020), Wang et al. (2020), Cai et al. (2020), Chan et al. (2020), Chen et al. (2020), Guan et al. (2020) estimate that the time from symptoms to hospitalization is between 5 and 12 days. The French Institut Pasteur relies on these estimates to announce a 5-day period from infection to onset of symptoms, followed by a 7-day period from symptoms to hospitalization.

2 As shown by Figure S2, available from Supplementary material B, this threshold correspond to 14 hospitalizations per 100, 000 inhabitants.

3 Supplementary material D show that results are robust to using a 10-day lag in lieu of a 11-day lag.

## Notes

* We are grateful to Antonio Brun Macipe, Jérémie Decalf, Romain Lutaud and Vincenzo Verardi for useful help and comments. Research on this project was financially supported by the Excellence of Science (EOS) Research project of FNRS O020918F.

### Competing Interest Statement

The authors have declared no competing interest.

### Funding Statement

Research on this project was financially supported by the Excellence of Science(EOS) Research project of FNRS O020918F.

### Author Declarations

IRB/oversight body approval/exemption does not apply.

